# Tracking and predicting the dynamics of HIV-1 epidemics in France using virus genomic data

**DOI:** 10.64898/2026.04.21.26351380

**Authors:** Louis Colliot, Vincent Garot, Paul Petit, Anna Zhukova, Marie-Laure Chaix, Laurence Meyer, Samuel Alizon, ANRS PRIMO Cohort Study

## Abstract

Understanding the dynamics of HIV epidemics is important to control them effectively. Classical methods that mainly rely on occurrence data are limited by the fact that an unknown part of the epidemic eludes sampling. Since the early 2000s, phylodynamic methods have enabled the estimation of key epidemiological parameters from virus genetic sequence data. These methods have the advantage of being less sensitive to partial sampling and to provide insights about epidemic history that even predates the first samples. In this study, we analysed 2,205 HIV sequences from the French ANRS PRIMO C06 cohort. We identified and were able to reconstruct the temporal dynamics of two large clades that represent the HIV-1 epidemics in the country. Using Bayesian phylodynamic inference models, we found that the first clade, from subtype B, originated in the end of 1970s, grew rapidly during the 80s before decreasing from 2000 to 2015 and stagnating since then. The second clade, from circulating recombinant form CRF02 AG, emerged and spread in the 80s, grew again in the early 2000s, before declining slightly. We also estimated key epidemiological parameters associated with each clade. Finally, using numerical simulations, we investigated prospective scenarios and assessed the possibility to meet the 2030 UNAIDS targets. This is one of the rare studies to analyse the HIV epidemic in France using molecular epidemiology methods. It highlights the value of routine HIV sequence data for studying past epidemic trends or designing public health policies.

**Author summary:** Despite huge progress in prophylaxis and treatment, HIV epidemics remain a major public health issue in most countries. Therefore, understanding, tracking, and predicting epidemic dynamics is essential to design optimal prevention and screening strategies. A strong limitation is that most methods rely on occurrence data and are very sensitive to the unsampled portion of the epidemic (also known as the ‘HIV hidden epidemic’). To address this issue, we take advantage of phylodynamics methods that rely on viral sequence data. Thanks to data from the ANRS Primo cohort, we identify two epidemics present in France since the early 1980s that exhibit consistent, but some times different, dynamics. By simulating future scenarios, we demonstrate that the UNAIDS goal to reduce new HIV infections by 90 % from 2010 by 2030 is uncertain, at least for one of the two epidemics we consider. This is one of the first studies to leverage phylodynamic methods to analyse the French HIV epidemic. It also highlights how routinely-generated genomics data can enable detailed analyses that facilitate the design of efficient public health policies.

## Introduction

Infections caused by the Human Immunodeficiency Virus (HIV) are among the first viruses for which sequencing data have been used to improve our understanding of their spread on a population scale [1]. This is because virus genetic sequence diversity can provide insights about an entire infected population, even if only a small fraction of this population is sampled [2]. The methods that allow such analyses, often referred to as phylodynamics, combine evolutionary biology and epidemiology to estimate key parameters that help to understand the epidemic [3]. HIV rapid evolution and routine sequencing make phylodynamics a suitable method for inferring the dynamics of the epidemic. This approach has already been applied to HIV epidemics in the UK [4–6], Latvia [7, 8], Poland [9], Portugal [10] and Switzerland [11–14] but remains underutilized in many other countries.

HIV incidence in France is currently estimated by mathematical models based on the mandatory declaration data associated with new HIV diagnoses. Recent analyses showed that incidence has been declining between 2012 and 2021, before stabilizing until 2024 [15]. Some epidemiological studies conducted in France used virus genetic data, e.g., to identify highly virulent variants (namely CRF 94 and 112 [16]), or to track variants resistant to antiretroviral treatments [17]. We also know that the proportion of HIV-1 subtype B among newly detected cases has been steadily decreasing over time [18]. Furthermore, one of the rare studies to perform phylogenetic inference revealed the existence of large transmission clusters in France [19]. However, we are unaware of any phylodynamic inference studies in the context of the French HIV epidemic, i.e., using genetic sequences to quantify epidemic spread. We conducted such an analysis using the nationwide ANRS PRIMO cohort data and demonstrated the insights it can bring to shape public health policies. More precisely, our main goal was to describe the past and ongoing structure of the HIV-1 epidemic in France, and to quantify its spread, especially by estimating key epidemiological parameters such as the temporal reproduction number *R*_*e*_ (i.e., the average expected number of secondary infections caused by a person living with the virus, at a certain date) or the duration of infection (i.e., the period during which an individual living with HIV can infect other people).

Before performing phylodynamic analysis on a set of sequences, we need to ensure that they emerged from a common introduction into the country of interest and belong to the same sub-epidemic. This step is necessary because even though the majority of HIV transmissions occur between people living in close geographical proximity, this is not always the case [20]. In other words, if we included all sequences sampled in France in a single analysis, we would obtain a representation of the worldwide epidemic, which is not our goal. To avoid this problem, we need to identify groups of sequences that are linked to the same virus introduction to the country and reflect transmission chains occurring within this country (here, France), as has already been done [10]. We hereafter refer to such groups of sequences as clades, a term from evolutionary biology that refers to groups of individuals formed by a common ancestor and all of its sampled descendants. After identifying some clades of interest, we applied phylodynamic methods to follow their historical dynamics and infer key epidemiological parameters. We then built on these estimates to explore several prospective scenarios regarding the effect of potential public health interventions on HIV spread. Finally, we built on prior knowledge about HIV epidemics, especially in France, to discuss the consistency of our results and identify future promising extensions of this study.

## Results

### PRIMO cohort population

During the study period (i.e., 1999 to 2024), subtype B was the most common subtype in the PRIMO cohort, with 62.9 % of the sequences available (Table 1). Among non-B subtypes, CRF02_AG was the most frequent with 15.9 % of the sequences. Note that 4.6 % of the sequences could not be subtyped by `jpHMM`. The proportion of subtype B sequences decreased over time from 77.6 % in 1999 to 28.6 % in 2024 (Fig S1). The median age at inclusion was 34 years for subtype CRF02_AG, 35 for B, 36 for C and others, 38.5 for unassigned, and 39 for subtype A. Men (87.7 %) were predominant in the cohort study and for all of the subtypes. ‘Men who have Sex with Men’ (MSM) represented the most frequent key population type in the cohort (68.8 %), followed by ‘Heterosexual’ (HTS, 19.5 %), ‘Injection Drug Users’ (IDU, 0.3 %), ‘Sex Workers’ (SW, 0.3 %), and ‘Suspected blood exposure’ (BLOO, 0.1 %).

**Table 1.**
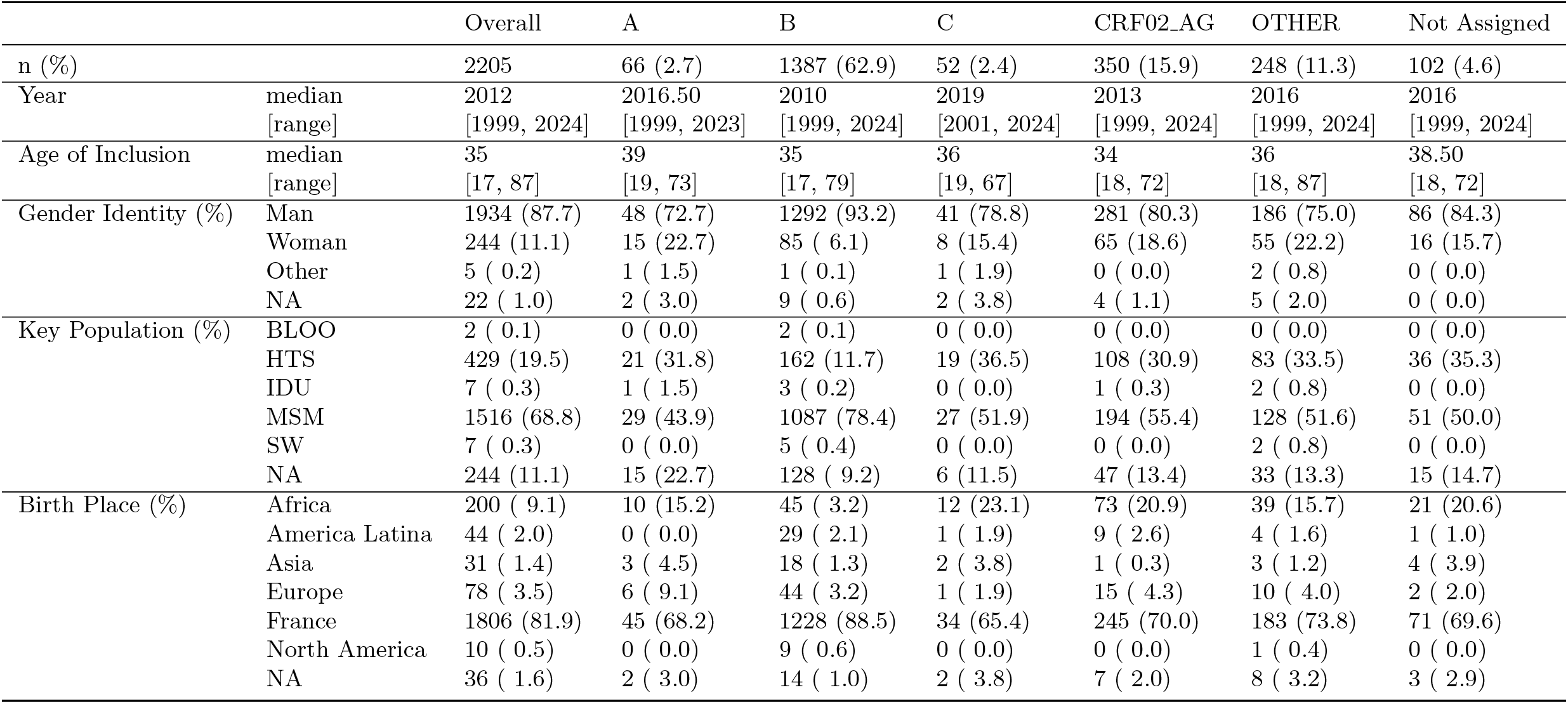
Characteristics of individuals included in the analysed subset of the PRIMO cohort, stratified by HIV-1 subtype. The population size is indicated as ‘*n*’ and also corresponds to the number of virus genetic sequences available. In the key populations, BLOO stands for ‘Suspected blood exposure’, HTS for ‘heterosexual’, IDU for ‘injecting drug users’, MSM for ‘men who have sex with men’, SW for ‘sex worker’, and NA for ‘not available’.

### Phylogeographic structure in the international context

To extract groups of sequences from the same part of the epidemic, we implemented a worldwide phylogeography approach (see the Methods on ‘Isolating French HIV clades’ and Fig 5), which we applied to the two main subtypes in PRIMO over the study period (B and CRF02_AG). In both cases, some sections/parts of the worldwide tree corresponded to specific geographic regions (i.e., had the same colour in Fig 1). The PRIMO sequences, in yellow, were distributed throughout the phylogenetic tree (Fig 1) but, both for subtype B and CRF02_AG, many clustered into large clades (identified as ‘Large Clade’ in Fig 1). These could be extracted automatically by assuming cut-off threshold (see the Mehtods) and yielded one clade with 129 sequences for subtype B (Fig S2) and another with 180 sequences for CRF02_AG (Fig S3).

**Fig 1.**
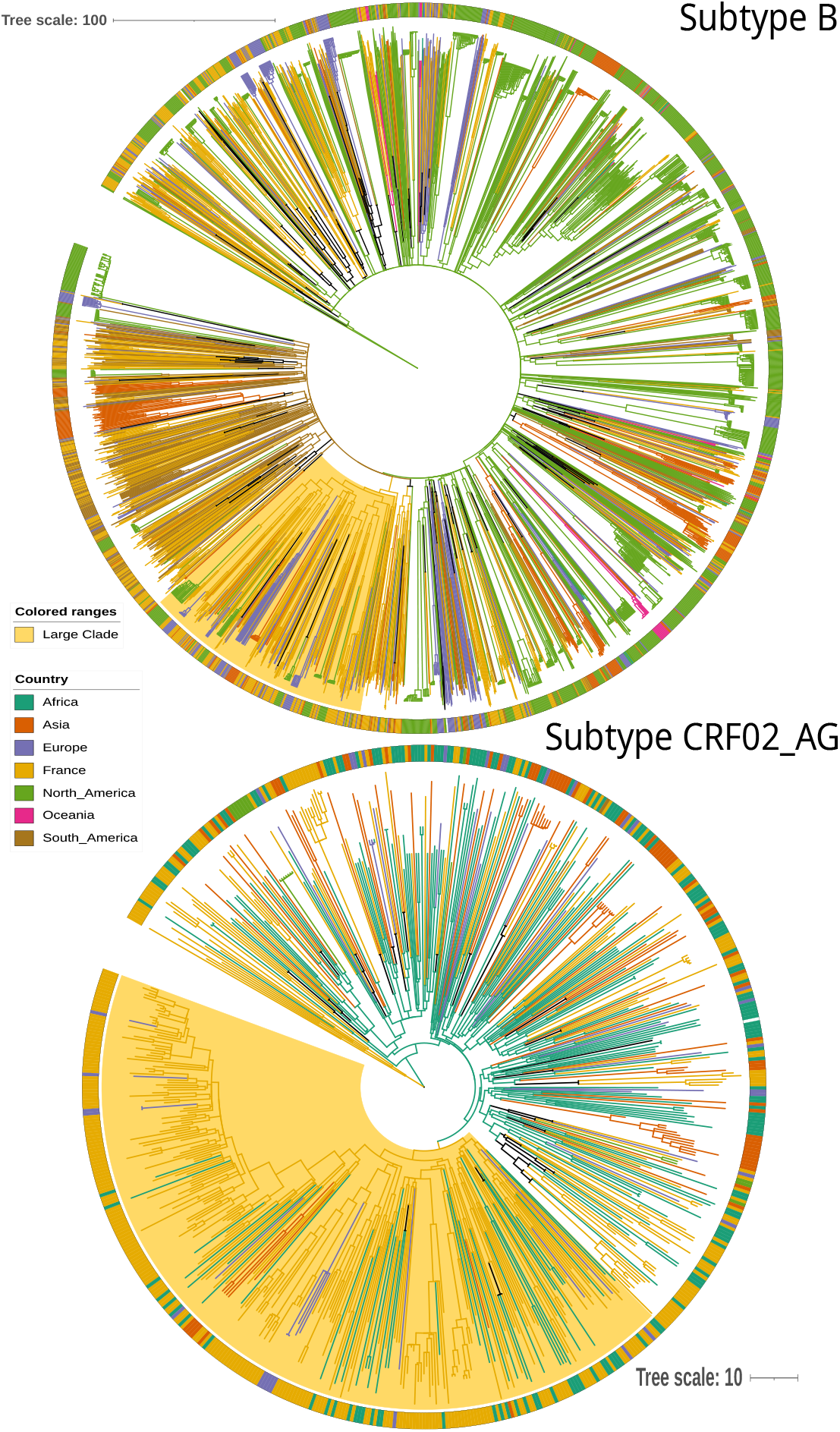
Phylogenies of the two dominant HIV subtypes in the PRIMO cohort in an international context. Sequences from the PRIMO cohort are in yellow and labelled as ‘France’. Sequences from countries other than France were obtained from the Los Alamos National Laboratory database and are coloured by continent. The largest PRIMO clade identified using `PastML` is indicated for each subtype with a light yellow area. The tree scale is in years and differs between the two trees.

Focusing on the patients from whom the sequences belonging to the large clade from subtype B originated revealed that they shared the same characteristics as most of the PRIMO cohort patients infected by HIV1-B, with the exception of a slight over-representation of men and MSMs (Table S1). Conversely, individuals corresponding to the CRF02_AG clade (Table S2) differed from other CRF02_AG-infected individuals, with an over-representation of men (93 % vs. 80 % for all CRF02_AG individuals) and MSMs (78 % vs. 55 % for all CRF02_AG individuals). Interestingly, the demographics of the individuals from the two clades were similar.

### Phylodynamics analysis of the large HIV-1 subtype B clade in PRIMO

Having identified large homogeneous clades, we performed phylodynamics analyses to infer past dynamics and estimate epidemiological parameters. We used two models, the Birth-Death SKYline model (BDSKY) and the COALescent SKYline (COALSKY), which offer complementary insights (see the Methods).

For the largest subtype B clade in the PRIMO cohort (5.9 % of all the sequences in PRIMO), the analysis using the BDSKY model dated the origin of the clade in 1977, with a 95 % high posterior density (HPD) from 1971 to 1982 (Fig 2C). The COASLKY model gave similar estimates with a 95 % HPD between 1972 and 1983 (Fig 2D).

**Fig 2.**
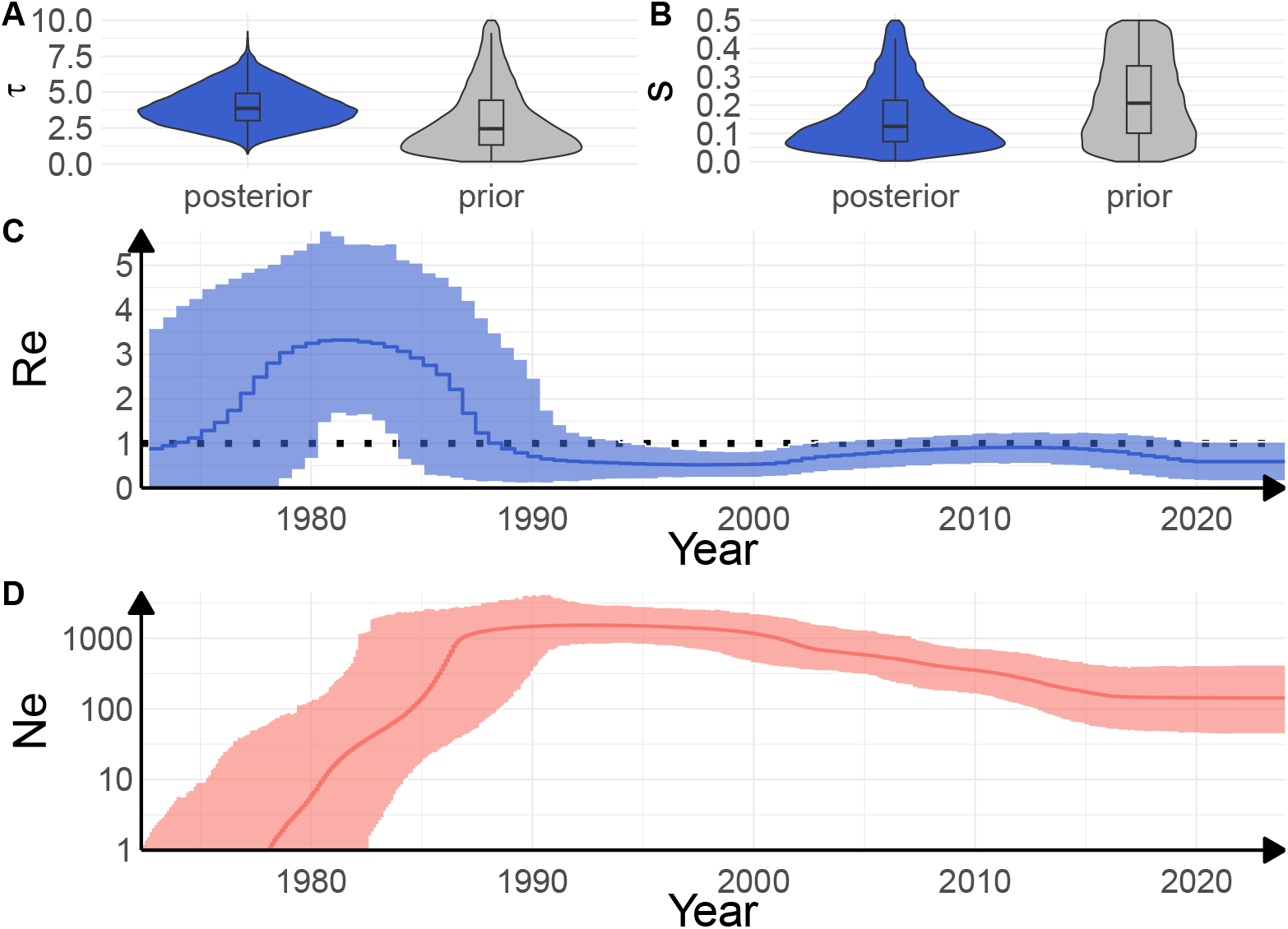
Phylodynamics analysis of the large subtype B clade (129 sequences) from the ANRS PRIMO cohort. Analyses assuming a birth-death skyline model are in blue, and those assuming a coalescent skyline model are in red. A) Posterior distributions of the infection duration (*τ*) and B) of the proportion of people who are sampled in the population (*s*). C) Temporal dynamics of the effective reproductive number (*R*_*e*_) and D) of the effective population size (*N*_*e*_). In A and B, boxes indicate the median, the lower bound Q1 at 25 %, and the upper bound Q3 at 75 %, the vertical lines indicate the range of values. In panels C and D, the areas indicate the 95 % highest posterior density (HPD), and the plain line the median of the posterior distribution.

The BDSKY model inferred a duration of infection (i.e., the time period during which an individual living with HIV is infectious, *τ*) of 3.9 years with an interquartile range (IQR) from 3.0 to 4.9 years (Fig 2A). Furthermore, among people living with HIV (PLHIV) who became non-infectious, the proportion for which this event is due to sampling and inclusion in the PRIMO database (*ρ*) was 12.5 % with an IQR between 7.1 and 21.7 (Fig 2B).

The same model found that the fastest growth of the epidemic occurred in the 1980s, with the median value of the effective reproductive number (*R*_*e*_) reaching 3.3 in 1983 (95 % HPD: 1.5–5.8). The median value remained above 1 until 1989, after which it stayed below 1 until our most recent estimates, with a 95 % HPD estimated to be between 0.2 and 1.0 in 2024.

These results are consistent with those obtained using the COALSKY model (Fig 2D), which estimates that the effective population size (*N*_*e*_) of the HIV-1B clade grew from the middle of the 1970s until 1991 to reach a maximum value of 1,489 individuals in 1991 (95 % HPD: 676–4310 individuals). After that, the population declined slowly until 2016, before plateauing until 2024, at which date its median was estimated to be 140 individuals (95 % HPD: 40–380).

### Phylodynamics analysis of the large HIV-1 CRF02_AG clade in PRIMO

The CRF02_AG clade contained 8.1 % (180) of all the sequences from the cohort. The BDSKY model dated its origin in 1980 (95 % HPD: 1975–1985; Fig 3C), which is consistent with the estimate from the COALSKY model with a median of 1979 and a 95 % HPD between 1973 and 1984 (Fig 3D).

**Fig 3.**
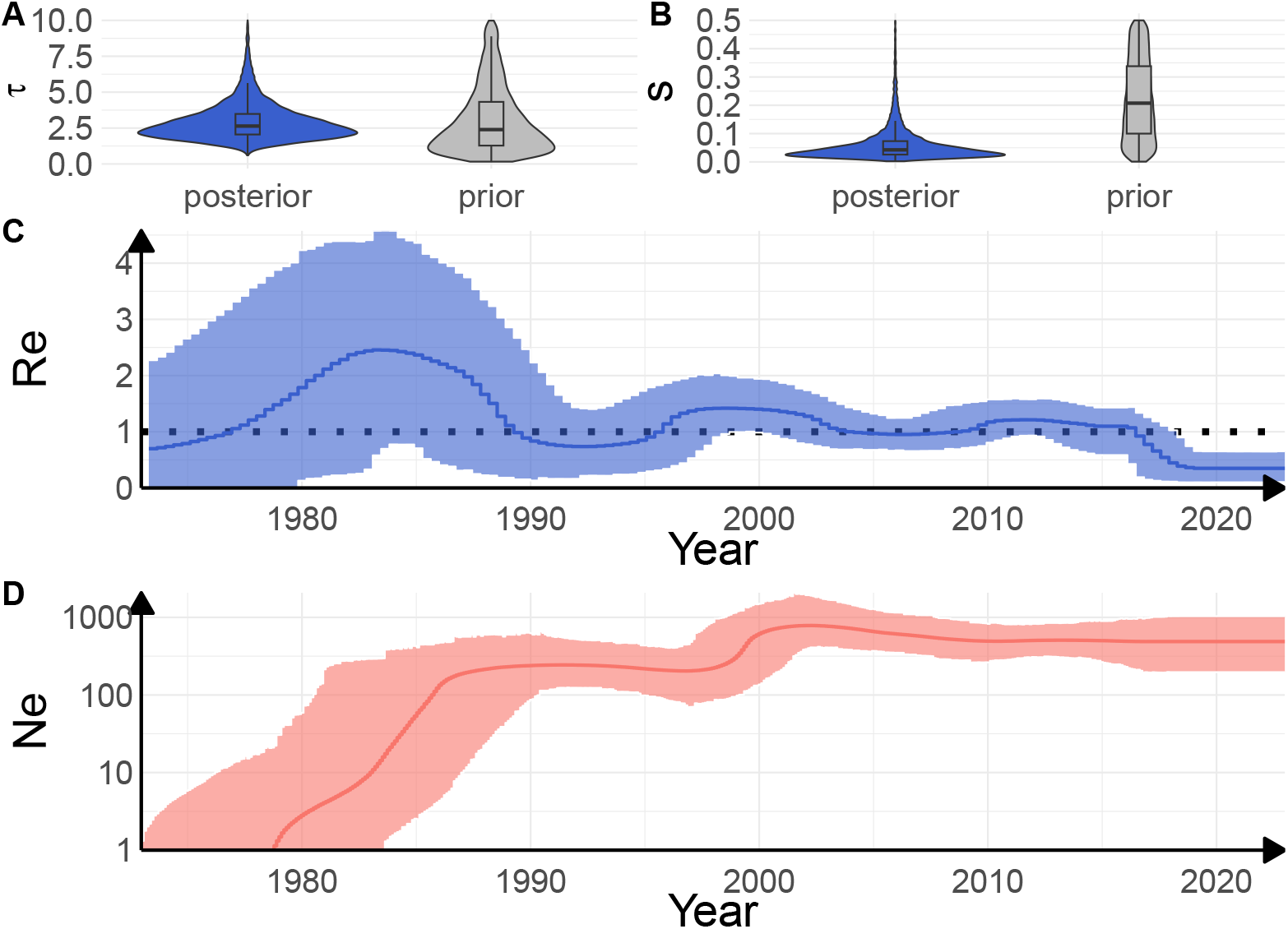
Phylodynamics analysis of the large subtype CRF02_AG clade (180 sequences) from the ANRS PRIMO cohort. See Fig 2 for details.

The duration of the infection (*τ*) was estimated by the BDSKY model to be of 2.6 years (IQR: 2.0–3.5; Fig 3A). The sampling rate (*ρ*) was estimated to be 4.2 % after 1999 (IQR: 2.5–7.4, Fig 3B).

The same model detected a growth of the epidemic in the end of 1970s, with an effective reproductive number (*R*_*e*_) that reached a maximum value of 2.5 (95 % HPD: 0.6–4.6) in 1984 (Fig 2C). The median *R*_*e*_ remained above 1 until 1990, after which it remained below this threshold until 1997. Contrarily to subtype B, periods of potential growth appear to have taken place in the early 2000 and, to a lesser extent in the early 2010. More recently, the clade size seems to have been strongly decreasing with estimated values of *R*_*e*_ in 2024 of 0.3 (95 % HPD: 0.1–0.7).

These results are consistent with those from the COALSKY model (Fig 2D), which estimates that the effective population size (*N*_*e*_) grew from the end of 1970s until 1990 to reach 231 individuals (95 % HPD: 47–599). The population size then remained stable until 1998, when there was another growth phase until 2003, when the population reached 786 individuals (95 % HPD: 410–1904). After that, the population remained stable until 2023, when it reached 488 individuals (95 % HPD: 197–1035).

### Prospective epidemiological scenarios

In essence, phylodynamics analyses infer past population dynamics. However, they do provide us with contemporary estimates for parameters of interest. We used the ones from the BDSKY model to simulate prospective scenarios after 2023. Assuming a birth-death model, we first simulated past dynamics from the inferred date of origin of each clade. These were very consistent with the effective population sizes estimates (*N*_*e*_) from the COASLKY model (Supplementary Fig S8). We then compared prospective scenarios, by different infection rates. In the neutral one, the parameter remains unchanged (in green in Fig 4); in the optimistic scenario, we assumed a 20% reduction in transmission (in orange); and in the pessimistic one, we assumed a 20% increase in transmission (in purple).

**Fig 4.**
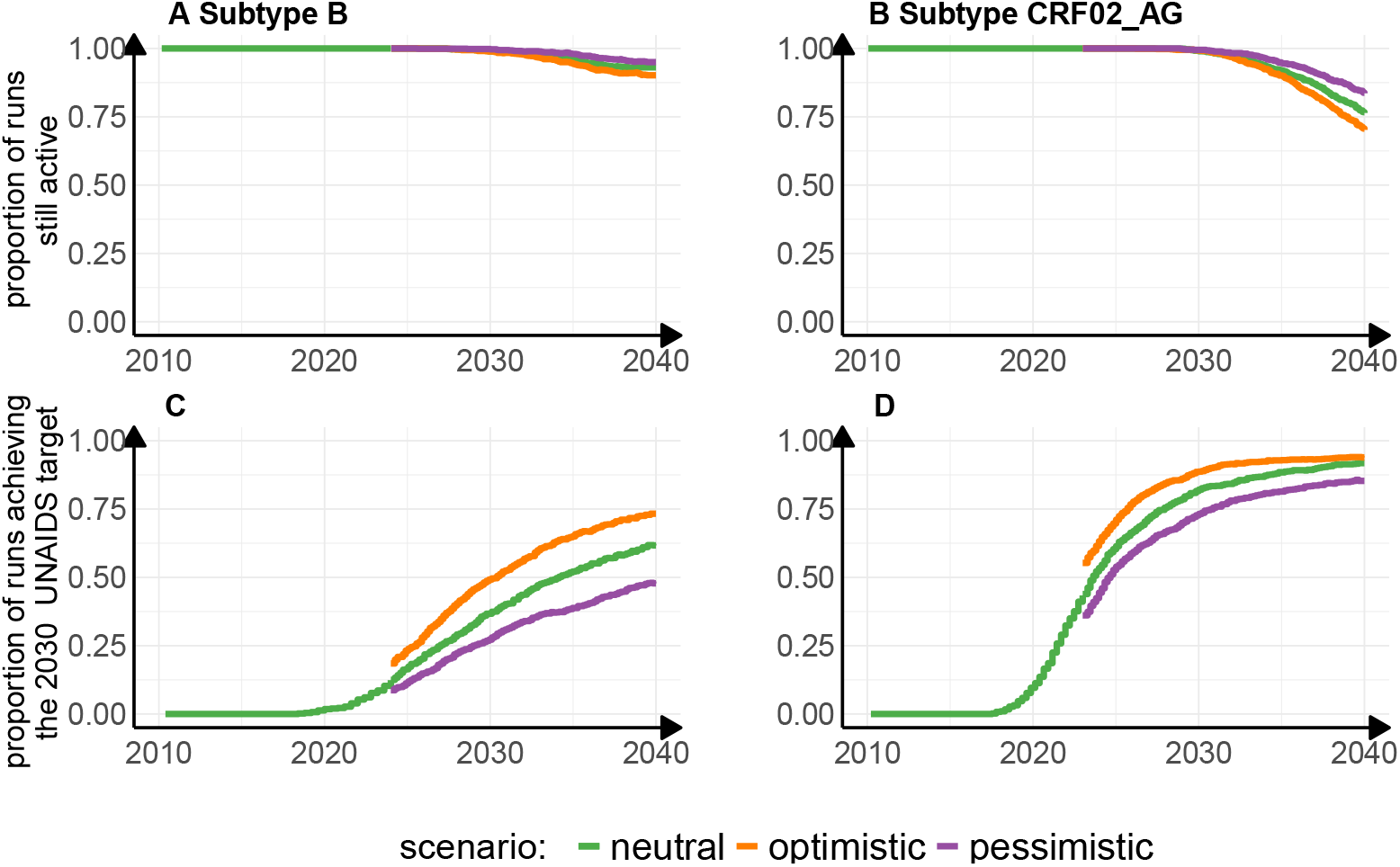
Simulated projection assuming neutral (in green), optimistic (−20 % transmission, in orange) and pessimistic (+20 % transmission, in purple) scenarios. A and B represent the proportion of simulations in which the main clade is still active. C and D represent the proportion of simulations that meet one of UNAIDS’ 2030 targets for the large PRIMO HIV subtype B (A) and CRF02_AG (B) clades. All three projections start from the latest BDSKY model estimates.

**Fig 5.**
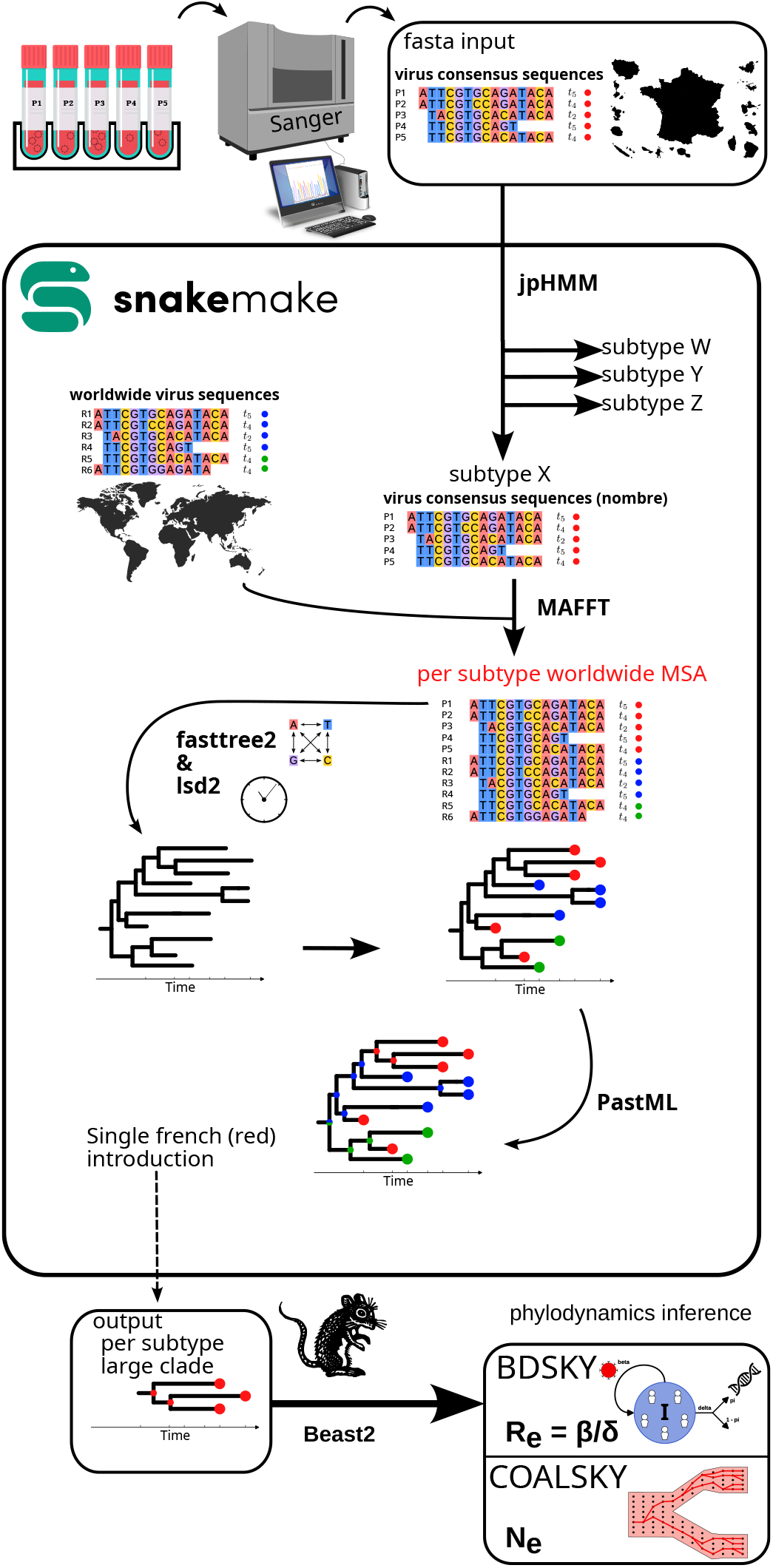
Structure of the bioinformatics pipeline for isolating homogeneous clades from a given HIV subtype. The input consists of a Fasta file containing HIV sequences of interest and a file with sampling dates. An initial subtyping step is performed using `jpHMM V0.1.4`. We then create a Fasta file for each subtype found in the data of interest. For each of these files, we add reference sequences from other countries using the LANL database. We then align the sequences of interest and the reference ones using `MAFFT V7.520`, before inferring a time-anchored tree using `FastTree V2.1.11`and `LSD V2.4.4`. Finally, we associate the sequences with their geographic region of origin and infer the spatial history of the clades with `PastML`. Homogeneous clades for a region of interest (in our case PRIMO sequences sampled in France) are extracted with `R`. The pipeline outputs a Fasta files and a table that contains the sampling dates for each clades. These can be used for phylodynamics inference with `Beast2 V2.4`.

For subtype B, the proportion of simulations in which the clade is still active in 2030 is above 0.98 in all three scenarios (Fig 4A). In 2040, this proportion decreases to 0.95 for the pessimistic scenario, 0.93 for the neutral one, and 0.90 for the optimistic one. For subtype CRF02_AG, the projection for 2030 is similar (Fig 4B). However, for 2040, these numbers decrease to 0.84 for the pessimistic scenario, 0.76 for the neutral one, and 0.70 for the optimistic one.

Focusing on one of the targets set by UNAIDS for 2030, namely achieving a 90 % reduction in HIV transmission compared to 2010, we find that for subtype B, the proportion of the simulations in which the main clade reaches these for HIV transmission is 0.28 for the pessimistic scenario, 0.37 for the neutral one, and 0.49 for the optimistic one (Fig 4C). In 2040, these numbers increase to 0.48 for the pessimistic scenario, 0.62 for the neutral one, and 0.73 for the optimistic one.

For the CRF02_AG subtype clade, the proportion of the simulations in which the main clade achieves the UNAID 2030 target in terms of reduction in HIV transmission is higher than in the subtype B clade with 0.73 in the pessimistic scenario, 0.82 in the neutral one, and 0.89 in the optimistic one (Fig 4D). At the 2040 horizon, these numbers are much higher with 0.85 for the pessimistic scenario, 0.92 for the neutral one, and 0.93 for the optimistic one. Since we estimate that the current size of the CRF02_AG large clade is larger than that of the subtype B large clade, this illustrates the importance of ongoing dynamics and the added value of phylodynamic insights.

## Discussion

HIV is a major public health issue worldwide, and quantifying its epidemic dynamics accurately timely remains challenging. In this context, the young field of phylodynamics is particularly useful because of its ability to leverage information from viral genomic data to better understand epidemic spread. Building on data from the ANRS PRIMO national cohort in France collected between 1999 and 2024, we demonstrate that these methods represent an asset to guide public health policies.

### PRIMO HIV epidemics are highly structured

That two virus genomes are more similar if they are sampled from the same geographical location indicates a spatially-structured epidemic. Focusing on the two main HIV subtypes in ANRS PRIMO cohort, we find that sequences can belong to different groups scattered across the international phylogenetic tree of a given subtype (Fig 1), which is consistent with multiple introductions of the subtype in France.

However, we also identify large homogeneous groups of sequences, which we refer to as clades, that are likely to share a common epidemiological history.

Within the ANRS PRIMO cohort, we identified two clades that belonged to two distinct subtypes. The clade sizes were similar (129 sequences for subtype B and 180 for CRF02_AG), even though the PRIMO database contains four times more subtype B than CRF02_AG sequences (1,387 *vs*. 350). One explanation of this difference is that there are fewer CRF02_AG reference sequences from other countries in Los Alamos National Laboratory database (in our final subtype B dataset, ANRS PRIMO sequences represent 7 % of all sequences, whereas this number increases to 30 % for the CRF02_AG one). This potential lower resolution for the CRF02_AG clade is also consistent with the differences in epidemiological dynamics that we further discuss below.

### Phylodynamics reveals consistent patterns among HIV subtypes

We used two complementary models to infer past epidemiological dynamics: the COALSKY is more phenomenological and mainly estimates variations in population sizes, whereas the BDSKY is more mechanistic and estimates population parameters. However, when using the BDSKY parameter estimates to simulate past population size dynamics, we obtained trajectories very consistent with the COALSKY model (Supplementary Fig S8).

The BDSKY model allows us to gain insights into the biology of the infections, for example by estimating the mean duration of infection. For both subtypes, the values that we estimated (3.0 to 4.9 years for subtype B and 2.0 to 3.5 years for subtype CRF02_AG) may seem short because a person infects others throughout the course of their infection. However, we also know that infectivity is higher during the first few weeks of infection, unless the person living with HIV is on effective antiretroviral treatment [21, 22]. Furthermore, in the underlying birth-death model, waiting times are exponentially distributed, implying that some individuals can remain infectious for a very long time. Finally, infectiousness can be reduced or suppressed thanks to behavioural changes or, more importantly, treatments.

The same model allows us to estimate the proportion of the end of infections that are due to sampling and inclusion in the PRIMO cohort, with 95 % HPD ranging from 7.1 to 21.7 % for subtype B and from 2.5 to 7.4 % for subtype CRF02_AG. According to the French national public health agency Santé Publique France (SPF), since 2012, approximately 25 % of the people who learned their HIV-positive status did so during primary infection [15]. This is larger than our estimates, but it is consistent with the fact that not all primary infections are included in the PRIMO cohort.

Regarding the historical dynamics, both the BDSKY and COALSKY models suggest that the B subtype clade epidemic grew from the late 1970s to the late 1980s. According to the BDSKY, there was a whole period of decrease from the mid-1990s to the early 2000s (the whole 95% HPD of the effective reproduction number, *R*_*e*_, was below 1). This could be due to the implementation of active antiretroviral combination. Subsequently, we estimate that *R*_*e*_ values did not exceed 1.3 (the upper bound of the 95 % HPD). We also identified a decrease in *R*_*e*_ in the years 2010s, which could be linked to the introduction of Treatment as Prevention (TasP) or Pre-Exposure Prophylaxis (PrEP) in the 2010s [23]. The COALSKY model has less resolution, which is expected given its simplicity (effective population size can only vary if there is a coalescence event in the phylogeny), but its trends are generally consistent with that of the BDSKY model.

The initial growth of the CRF02_AG subtype clade appears to be contemporary to that of the subtype B clade and in both cases we observe a stagnation of the epidemic in the 1990s. However, for the CRF02_AG clade, and for both the BDSKY and COALSKY models, we detect a second growth at the end of the 1990s that contrasts with the declining dynamics of the subtype B clade. A closer look at the CRF02_AG phylogenetic tree (Fig S5) helps to understand this epidemic rebound. Indeed, the clade of interest appears to contain a subclade that emerged in the early 1990s. One potential explanation is that there was at least one other introduction event and that we failed to separate our large clade into smaller clades because of the lower number of reference genomes for this subtype. Another possibility is that there was a change on the host side at the end of the 1990s and that this epidemic was refueled by another population. Sociodemographic information shown in Table S2 tends to support this hypothesis because 87.5 % of the individuals in the new small subclade were identified as MSM compared to 68.5 % of the individuals in the remaining of the large clade. More generally, the sociodemography of the small subclade was very similar to that of the large clade of subtype B.

Finally, for both clades, the most recent estimates from the BDSKY and COALSKY models diverge, with *R*_*e*_ indicating a declining epidemic whereas *N*_*e*_ a stagnating one. A known explanation is that the COALSKY model assigns a constant effective population size over a period of time. The problem is that, as shown by Fig S4, Fig S5 and Fig S6, there are few coalescent events in recent time periods. Therefore, the constraints in terms of tree topology make it impossible to update the effective population size estimates.

### Meeting the UNAIDS 2030 objectives is uncertain

Our projections based on contemporary estimates from the phylodynamics inference suggest that by the year 2030, the two main clades we identified in the ANRS PRIMO cohort exhibit contrasting trends in terms regarding our ability to reduce new HIV infections by 90 % compared to 2010. Even in the best-case scenario, the B clade has less than 50 % chances of reaching this UNAIDS target, whereas for the CRF02_AG clade, even in the worst-case scenario, this probability is greater than 70 %. This difference reflects the latest dynamics observed in the *R*_*e*_ for both clades (Fig 2 and Fig 3). After 2030, for the CRF02_AG clade, the dynamics appear to slow down, which is not the case for the subtype B clade. By 2040, almost all simulations have reached the target for the CRF02_AG subtype (85.3 % for the pessimistic scenario). The results for subtype B are less favourable, but still range from 48 % in the pessimistic scenario to 73 % in the optimistic one. Nevertheless, for both clades and across all scenarios, it is quite uncertain whether these two populations will be inactive even by 2040. This implies that even if the UNAIDS target is met, efforts must continue, lest the epidemic spiral out of control again in these two clades.

This underlines the potential impact of public health policies since the variations in transmission rate (*β*) we simulated are consistent with field estimates regarding the effect of TasP and PrEP [24] and the fact that PrEP remains underused in France in some key populations [25]. Nevertheless, according to our estimates, even with increased use of PrEP, meeting the aforementioned 2030 targets seems uncertain.

### Future potential model refinement

A limitation of our study is that we assumed a simple underlying epidemiological model with only one type of individual. Such level of detail does not allow us to capture variations in social, demographical, and epidemiological heterogeneity. However, this limitation is mitigated by the fact that we focus on homogeneous clades, both in terms of spatio-temporal aspects and of socio-demographic characteristics (as illustrated by Tables and). This means that including a larger fraction of the PRIMO sequences into the analysis would require more detailed models, e.g., with multiple host types.

Another limitation has to do with our assumption that only the transmission rate (*β*) can vary over time, while the duration of infectiousness (*τ*) and the proportion of sampling (*ρ*) remain constant. In reality, all three are likely to have varied over the last few decades. However, due to model’s identifiability issues [26], we are unable to obtain satisfactory posterior densities to infer a clear dynamics of the epidemic without making such simplifying assumptions. This is also why we only reported variations in reproduction number (*R*_*e*_), which integrates both components. Increasing the sampling proportion, e.g., by focusing on a specific densely-sampled region in France, could help to alleviate this limitation.

Finally, only a fraction of the available sequences was used in the final phylodynamic analyses. This choice was made to ensure the consistency between the set of sequences analysed and the underlying assumptions of the phylodynamics models. It also allowed us to highlight the contrasted dynamics of two persistent and active transmission networks, while demonstrating that it is potentially misleading to refer to a single ‘global’ epidemic in a country. As mentioned above, a possibility could be to use more detailed phylodynamics models to capture epidemic migration between countries.

However, this would require analysing much more sequences (that from the country of interest along with a worldwide reference dataset), which would quickly generate computational hurdles and most likely require to separate the phylogenetic inference and the phylodynamics inference steps [27]. Another possibility could be to study all the clades from a given subtype, even the smallest ones, independently and merge the estimates but this could require to simplify the model even further, especially with respect to temporal variations.

### Perspectives

Using only a small fraction of the sequence data routinely collected in France, we show that it is possible to identify large clades that reflect the past and current status of the epidemic. We also show that phylodynamic methods complement traditional occurrence-based ones because they readily allow to focus on specific epidemics (instead of analysing the epidemic as a whole) and give access to some parameters that are difficult to infer, such as infection duration. Phylodynamic methods also open a window on the history of the epidemic, since they can even provide insights about time periods that pre-date the first sample in the dataset. Although we created a dedicated pipeline to process HIV sequence data, additional work is required to further demonstrate the feasibility of integrating such phylodynamics methods in routine surveillance protocols. On a more conceptual level, this is one of the first studies to use only sequences from individuals detected during primary infection for phylodynamics analyses and it will be interesting for future studies to investigate to what extent this specificity may increase the accuracy of the estimates.

## Materials and methods

### The ANRS PRIMO Cohort

As further described in other articles [17, 28], the ANRS PRIMO cohort has the particularity of including only PLHIV diagnosed during an acute or early phase of their infection. It has been running since 1996 and we used the virus `pol` gene sequence and the year of sampling for each participant included during the period 1999–2024. We also used socio-demographic data such as the gender (man, woman, other and NA for ‘not available’), the age, the HIV key population (heterosexual, men who have sex with men, sex worker, injecting drug users, suspected blood exposure, and NA), and birth place.

### Isolating French HIV clades

We developed a pipeline (https://gitlab.college-de-france.fr/lcolliot/isoclade) to identify clade, i.e., groups of HIV sequences from a given country (in this case, France) that share a common evolutionary history. We first generated a dataset for a given HIV subtype with focal sequences. For each sequence, the subtype was estimated using `jpHMM V0.1.4` [29]. To provide a global context, reference sequences of the same subtype from other countries and with known sampling dates listed in the Los Alamos National Laboratory database (LANL, https://www.hiv.lanl.gov/content/sequence/NEWALIGN/align.html) were added to each subtype dataset (Supplementary File). We then aligned the ‘focal’ and ‘reference’ sequences from our new dataset using `MAFFT V7.520` [30] with default parameters. For the resulting multiple sequence alignment (MSA), a phylogeny was inferred using `FastTree V2.1.11` [31] under a general time-reversible (GTR) model with a discrete gamma distribution to model inter-site rate variation and 1000 ultrafast-bootstraps. In the inferred phylogenetic tree, distances are given in number of substitutions but these can also be expressed in number of years by leveraging information from the sampling dates. We used `LSD V2.4.4` [32] to perform this time calibration and also to remove temporal outliers. Finally, we used `PastML` [33] to infer past geographic spread. This step associates each internal node with a marginal probability that it originated from each of the geographical locations in the dataset. To define a clade, we used a threshold of marginal probability *>* 0.90 for its internal node geographical locations, and kept homogeneous clades of more than 50 sequences. The trees resulting from the `Pastml` analyses shown in the manuscript were generated with `iTOL` [34].

### Epidemiological dynamics inference using Beast2

Having identified phylogenetic clades associated with a part of the focal HIV epidemic, we assumed that the parameters we were going to estimate for them were homogeneous. It means that we assume that the individuals in the population are relatively similar according to the epidemic. We analysed each of these separately using two classes of phylodynamic models, namely birth-death (BD) and coalescent models, both of which are implemented in the software package `Beast2 V2.4` [35].

We used the Coalescent Bayesian Skyline (COASLKY) model [36] to estimate temporal variations in HIV effective population size. This variable is a delicate concept to handle, especially in the case of HIV sequence data [37]. It is based on the amount of observed genetic diversity and can be defined as the population size carrying this diversity would need to have in order for it to be consistent with a Wright-Fisher population genetics model [38]. Here, the population refers to the part of the epidemic that has been sampled via the homogeneous clade of interest. Note that an underlying assumption of this coalescent model is that the proportion of the population sampled is negligible, which is appropriate for our dataset.

We also used the birth-death skyline (BDSKY) model [12] to estimate a transmission rate (*β*), a rate at which individuals become non-infectious (*δ*), and a sampling proportion (*s*). The latter corresponds to the proportion of end of infectivity events that result from individuals being sampled and included in our dataset. The effective reproduction rate (*R*_*e*_) is defined by *β/δ* and represents the average expected number of secondary infections caused by a person living with the virus, at a given time. The duration of infection (*τ*) corresponds to the period during which an individual infects other people and is equal to 1*/δ*. The infectious period can end for several reasons, one of which being the sampling because we assume that once an individual living with HIV knows their status, they change their behaviour and no longer transmit the virus. The BDSKY model assumes that the reproduction number *R*_*e*_ is piecewise constant with 10 temporal windows, that *τ* is constant, and that *s* is zero before 1999, the date of the first sampling event, and constant after that. The sampling proportion corresponds to the sequences present in the PRIMO cohort, which is why we assume sampling was negligible before 1999 although sampling did take place before the cohort was established. Compared to the coalescent model, the birth-death one offers more mechanistic insights by disentangling the transmission rate and the duration of infection [39].

### Prospective scenario modelling

To explore the future dynamics of the HIV epidemic, we used the estimates from the BDSKY model (*β* and *δ*) to simulate prospective scenarios. First, we simulated the number of PLHIV in our clades of interest, from the start of each epidemic until 2023, assuming a birth-death model. This was done using a Gillespie algorithm [40] in `R`. The resulting epidemiological trajectories were compared to the *N*_*e*_ estimated with the COALSKY model to check for consistency in population sizes.

To simulate prospective part of the scenarios, we used the last time point of the previous simulation as a starting point and extended it by varying the last value of *β* to simulate the effect of interventions, *e*.*g*., expanding or restricting access to pre-exposure prophylaxis (PreP). We build on the results from the work from Boyle et al. [24], who estimate that in Canada, between 2016 and 2023, 20 % of the infections could have been prevented thanks to PreP. Overall, from 2023, we compared three scenarios: a neutral one with the same transmission rate (*β*), an optimistic one with a 20 % decrease in *β*, and a pessimistic one with a 20 % increase. We simulated 1,000 replicates for each scenario. Using these simulations, we calculated the proportion of simulations that remained active (i.e., with at least one PLHIV) over time, as well as the proportion that achieved the infection reduction target set by UNAIDS for 2030. This target consists in a 90 % reduction in the number of new infections compared to 2010. To calculate the number of new infections at any given time point, we multiply the number of PLVHIV by the temporal reproduction number value (*R*_*e*_).

## Data Availability

All data produced in the present study are available upon reasonable request to the authors

## Supporting information

**Table S1 Characteristics of the individuals included in the PRIMO cohort with a focus on the large clade from subtype B**. See Table 1 for additional details.

**Table S2 Characteristics of the individuals included in the PRIMO cohort with a focus on the large clade and the subclade from CRF02_AG**. See Table 1 for additional details.

**Fig S1 Samples proportion by subtype over time**

**Fig S2 Pastml’s result for subtype B:** The Fig represents the evolutionary history of a trait, in this case geographical location (PRIMO in a global context). The circles represent the state, and the arrows represent the change in state of the trait.

**Fig S3 Pastml results for subtype CRF02_AG showing PRIMO in a global context**. The graph represents the evolutionary history of a trait, in this case geographical location. The circles represent the states and the arrows the change in state of the trait (here the geographical location).

**Fig S4 Phylogeny of the PRIMO large B clade**.

**Fig S5 Phylogeny of the PRIMO large CRF02_AG clade. Fig S6 Phylogeny of the PRIMO CRF02_AG subclade**.

**Fig S7 Phylodynamics of the subclade subtype CRF02_AG clade (96 individuals) from the PRIMO cohort**. See Fig 2 for details.

**Fig S8 Comparison between birth-death simulations and the estimations from the COALSKY model for large PRIMO clades**.

## Acknowledgments

LC is supported by a PhD fellowship 13643 from Sidaction. PP is supported by a PhD fellowship 20275 from Sidaction. This work was also partly supported by the by the Agence Nationale de la Recherche for the DEELOGENY project (ANR-23-CE45-0027). VG is supported by a PhD fellowship by the Agence Nationale de la Recherche for the DEELOGENY project (ANR-23-CE45-0027). We thank all the patients who are participating in the ANRS PRIMO cohort and the ANRS PRIMO Cohort scientific committee (V Avettand-Fenoël, F Barin, C Bourgeois, ML Chaix, A Cheret, S Couffin-Cadiergues, JF Delfraissy, A Essat, H Fischer, C Goujard, C Lascoux-Combe, C Lecuroux, L Marchand, L Meyer, C Rouzioux, A Saez-Cirion, R Seng). We also thank all physicians participating to the ANRS PRIMO Cohort study.

